# The impact of adherence to the Australian social media ban on adolescents’ psychological distress and daily functioning: an emulated trial

**DOI:** 10.64898/2026.11.24.26368812

**Authors:** Stéphanie Baggio, Mado Gautier, Marcela Radunz, Christina R. Galanis, Matthew W. R. Stevens, Diana Khanna, Joël Billieux, Vladan Starcevic, Katajun Lindenberg, Tracey Wade, Daniel L. King

## Abstract

**Background:** Australia was the first country to adopt a national minimum age requirement for social media use. This policy has been widely debated, and evidence is only just starting to emerge about its impacts on adolescents. This study examined the effect of the social media ban on psychological distress, health-related quality of life (HRQoL), and the impact on daily life and academic self-efficacy.

**Methods:** We conducted an emulated trial using follow-up data collected three months after the ban implementation from a two-wave survey of South Australian high school students aged 12 to 15 (N=551). Exposure was defined as adherence to the ban, which was assessed in two ways: strictly (self-reported adherence combined with reported social media behaviours), and less strictly (self-report only). The primary outcome was the Revised Children’s Anxiety and Depression Scale (RCADS-11). Secondary outcomes included RCADS-11 impact subscore; Child Health Utility 9 Dimensions (CHU9D) utility score (HRQoL); score of CHU9D activity-related items, which assessed daily functioning; and the academic self-efficacy subscale of the Self-Efficacy Questionnaire for Children (SEQ-C). Adjusted regression models were performed using inverse probability weighting to account for group imbalance and attrition.

**Findings:** Using the strict definition, 14.8% of adolescents adhered to the ban, compared to 41.5% under the less stringent definition. Adherence had no significant effect on the RCADS-11 total score or impact subscore (p ≥ .321). Those who adhered to the ban had significantly higher HRQoL, better daily functioning and higher academic self-efficacy (p ≤ .033). Overall, effects were small in magnitude. No significant effects were observed under the less stringent definition of adherence (p ≥ .244).

**Interpretation:** Strict adherence to the social media ban was associated with minor short-term improvements in HRQoL and daily functioning, but not psychological distress.

**Funding:** This research was supported by a Channel 7 Children’s Research Foundation grant.

**Research in context:** *Evidence before this study:* On 1 November 2026, we searched the PubMed database from its inception with no language restrictions, using the terms (“social media” AND “ban” AND “Australia”) OR (“Social Media Minimum Age Act”). We included studies that reported empirical data on Australia’s social media minimum age policy. Of the 47 records identified, only one observational study reported empirical findings on the ban. Using manual search, we added another study. Both studies found that adolescents continued to use social media after the ban was implemented. The second study showed that reducing social media by ≥25% was associated with small decreases in depression and anxiety. Outcomes related to daily functioning were not assessed. Both studies had high risk of bias due to their observational design and sample size/dropout rate.

*Added value of this study:* To our knowledge, this was the first study to use an emulated trial framework to address confounding and attrition in order to estimate the effect of adherence to a national social media ban on adolescents’ psychological distress, health-related quality of life (HRQoL), and daily functioning. Only 14.8% of adolescents strictly adhered to the ban three months after its implementation, compared to 41.5% who reported doing so. Strict adherence was not associated with lower psychological distress, but was associated with small improvements in HRQoL, daily functioning, and academic self-efficacy. No effects were observed when adherence was based on self-report alone.

*Implications of all the available evidence:* The benefits of the ban appeared to be limited to adolescents who fully disengaged from social media, and the low level of adherence was likely to reduce its impact at a population level. Policies should prioritise enforcement measures that hold platforms accountable, alongside psychoeducation for young people and parents. Future evaluations should include daily functioning as a key outcome and use objective measures of adherence. They should also include longer follow-ups and be powered to detect small effects.

## Introduction

Social media has become an integral part of daily life. The technology enables social connectivity, self-expression, and sharing ideas and content across networks (1). However, healthy or adaptive social media engagement can become habitual and develop into an excessive pattern of use, with some individuals developing compulsive use and addiction-like symptoms (2). Problematic use of social media is linked to reduced mental and physical well-being, with studies highlighting these negative consequences already in young users (1–4). Adolescents are more vulnerable to developing problematic social media use due to developmental factors such as greater impulsivity and lower emotion regulation, immature decision-making skills, and sensitivity to peer influences (5).

The widespread use of social media among adolescents has become a global concern and the focus of various public health interventions motivated by the precautionary principle. Policymakers and other stakeholders (e.g., parents, teachers) are keen to know if these countermeasures have produced meaningful benefits for the most vulnerable population of users. This is particularly salient for Australia, the nation that introduced the first age restriction policy for social media (affecting those under 16 years) in December 2025. Currently, the scientific evidence on the effectiveness of these interventions is limited due to the recency of these developments (6).

Two recent systematic reviews and meta-analyses focusing on experimental designs reported that social media restrictions could have small positive effects on well-being (7, 8). Many studies included in these reviews have limitations. They have relied on convenience samples of adults, including volunteers, and had high dropout rates and only short-term interventions. Furthermore, these restrictions are often partial, allowing reduced social media use or social media use at certain times of the day. Therefore, their results cannot inform us about the potential benefits of nationwide bans targeting adolescents. Further, very few studies have reported empirical findings on the recent Australian social media ban. Barnes et al. (9) examined adherence to the ban, reporting that 85% of adolescents continued to use social media after the ban. To our knowledge, few studies have evaluated the impact of the Australian ban on psychological distress and daily functioning. Radunz et al. (10) found evidence of small benefits of a minimum-age social media policy on adolescent mental health.

Daily functioning refers to a person’s ability to carry out the activities and fulfil the roles expected of them. For adolescents, this encompasses family and home life, friendships, classroom learning and homework, leisure activities, and maintaining daily routines (e.g., getting enough sleep, staying active, and eating healthily) (11). Problematic social media use may be associated with impaired daily functioning as it can interfere with everyday activities, schooling, and sleep (2, 4, 7). However, experimental evaluations of social media restriction have almost exclusively focused on well-being and mental health. To overcome these limitations and advance our understanding in this new area of public health, this study employed an emulated trial framework to evaluate the impact of the Australian social media ban on psychological distress and daily functioning. The primary outcome was psychological distress. Secondary outcomes included health-related quality of life (HRQoL), impact on daily life, and academic self-efficacy.

## Methods

### Design

This study used data from a cohort survey that evaluated the impact of the Australian social media ban to emulate a trial using inverse probability weighting. The ban set a minimum age of 16 to use social media platforms such as Facebook, Instagram, Snapchat, TikTok, and YouTube. Data were collected at two time points: in November 2025 (baseline, before the ban) and in March 2026 (follow-up, three months after the ban was implemented); the design and data have been described in detail elsewhere (10).

### Setting and participants

Data were collected from 13 out of 57 invited public high schools in South Australia. Participants from Years 7 to 12 (aged 12 to 18) were invited to take part during class time. The study included participants who were aged 12 to 15 at the time of data collection. Those aged 16 or over were excluded, as the ban does not apply to them. Adolescents who did not own any social media account were also excluded.

The study was approved by the Flinders University Human Research Ethics Committee (project number 8751) and the Department of Education and Child Development Ethics Committee (project number 049-2025). Informed consent was obtained from students via an opt-in procedure and from parents via an opt-out procedure. The project was pre-registered on the Open Science Framework (see: https://osf.io/q2tb9).

### Adherence

Adherence to the ban was defined as a binary variable (yes/no), assessed at follow-up. The “yes” category included participants who reported never having had a social media account, as well as those who no longer had any at follow-up. The “no” category included participants who reported continued use of social media accounts at follow-up, or who reported being over 16 years of age, even if their self-reported age was lower than 16 years (n = 9). To further correctly assess adherence, the following self-reported follow-up behaviours were considered as non-adherence: reporting sharing/posting on social media and indicating regular use of social media accounts targeted by the ban in the previous three months (not including YouTube, which is easily accessible without an account, and ’other’ platforms, as we could not tell whether they required an account or not). These variables were used to derive a strict definition of adherence. A less stringent definition, based solely on self-reported adherence to the ban, was used for sensitivity analysis.

### Outcomes

*Revised Children’s Anxiety and Depression Scale (RCADS-11).* The RCADS-11 at follow-up was used as the primary outcome to assess psychological distress (12). This scale includes eleven items on a four-point Likert scale scored from 0 to 3, focusing on depressive (five items) and anxiety (six items) symptoms. A sum score was calculated (range 0–33), with a higher score indicating higher psychological distress. Additionally, two items assessed functional impact using a 4-point scale scored from 0 to 3 (“*How much do these difficulties upset or distress you?*” and “*How much do these difficulties get in the way of your everyday life in school?*”). We used the sum score (range 0–6) at follow-up as a secondary outcome to evaluate the effect of depressive and anxiety symptoms on daily life.

*Child Health Utility 9 Dimensions (CHU9D)*. The CHU9D at follow-up was used as a secondary outcome. It assesses HRQoL and is used in economic evaluations of healthcare interventions (13). It comprises nine dimensions: worried, sad, pain, tired, annoyed, schoolwork/homework, sleep, daily routine, and able to join in activities, each with 5 severity levels (1 = ‘no problems’ to 5 = ‘most severe’) and asks respondents about their HRQoL today. The CHU9D responses can be converted into a utility score anchored at 0 (dead) and 1 (full health), with negative values representing states worse than dead. We used the Australian adolescent-specific algorithm to calculate the utility score, with a minimum score of -0.1059 (14). Additionally, we used a sum score of three items evaluating daily life functioning (activity-related items: schoolwork/homework, daily routine, and able to join in activities, range 3–15) at follow-up, where a higher score indicates worse functioning.

### Self-Efficacy Questionnaire for Children (SEQ-C)

The academic subscale of the SEQ-C at follow-up was used to assess functioning related to academic achievement (15). This subscale comprises seven questions, each assessed on a 5-point Likert scale, scored from 1 to 5. Examples of these questions include “*How well do you succeed in finishing all your homework every day?*” or “*How well can you pay attention during every class?*”. A sum score was calculated (range 7–35), with higher scores indicating greater academic self-efficacy.

### Covariates

Potential confounding factors were identified through a literature review. Where possible, studies focusing on the moderators of the effects of social media use were used; otherwise, studies focusing on the effects of a mobile phone ban were used (8, 16, 17). Baseline covariates included: age (in years); gender (girl, boy or non-binary); main language spoken at home (English or other) as a proxy for cultural background; socio-economic status, as assessed by the Socio-Economic Indexes for Areas (SEIFA) score; social media use, as assessed by the number of hours spent on social media each week (17); bullying, as assessed by the Bullying and Cyberbullying Scale for Adolescents (BCS-A), a scale assessing offline (8 questions) and online (5 questions) victimisation using a 5-point Likert scale, scored from 0 to 4 (18); and an indicator of mental health, as assessed by the baseline RCADS-11 total score. The baseline level of each outcome, including CHU9D and academic self-efficacy, was also included in the models.

### Statistical analyses

We performed a sensitivity power analysis using G*Power 3.1 to determine the minimum effect size that could be detected. Using a two-tailed independent t-test with α=.05, 1-β=.80 and sample sizes of 471 and 80 in the non-adherer and adherer groups, we could detect moderate effect sizes (Cohen’s d=0.34).

Analyses were performed on the follow-up sample using baseline covariates to calculate inverse probability weights (see below). In a preliminary step, we calculated descriptive statistics (means and standard deviations, percentages and n) for the sample and each group, along with 95% confidence intervals (CI) for adherence. The analyses then combine three methods of addressing baseline missing values, attrition, and potential group imbalance to emulate the trial. First, missing values on baseline covariates were imputed using imputation by chained equations. The imputation models were customised for each variable’s scale. We used augmented logistic or multinomial logistic regressions for categorical variables and truncated linear regressions for continuous variables. A single imputed dataset (m=1) was generated. We selected this approach because full multiple imputation failed on our dataset, resulting in instability of the multi-stage weights across imputations and rendering Rubin’s rules inapplicable.

Second, the groups (adherers and non-adherers) were balanced to mimic a randomised controlled trial by using inverse probability of treatment weighting (IPTW). This method creates a pseudo-population in which exposure is not associated with covariates (19). Third, since there were many non-completers at follow-up, inverse probability of attrition weight (IPAW) was used to better account for selection bias due to attrition. Baseline covariates were used as predictors in the IPTW/IPAW models. The analyses were performed using stabilised, truncated IPTW/IPAW. We report the SMD between groups after IPTW/IPAW, with an SMD ≤. 25 indicating successful balancing (20, 21). Fourth, we tested the impact of adherence to the social media ban using linear regression models with IPTW/IPAW, adjusting for baseline covariates and accounting for potential clustering effects using robust standard errors.

We also ran sensitivity analyses using alternative models. To account for the bounded nature of the CHU9D utility score and its ceiling effect (n=165 with a score of 1 at baseline and n=68 with a score of 1 at follow-up), we employed a fractional logit model (22). Negative values were recoded as zero, since this model cannot handle negative values (n=12 at baseline, n=6 at follow-up). As the score of CHU9D activity-related items was positively skewed, we conducted a sensitivity analysis using negative binomial regression. The results were consistent with those reported in the Results section (statistical significance, magnitude of the associations). Finally, all analyses were also performed using a less stringent definition of adherence, with the results reported in the supplementary material. All analyses were performed using Stata 19.5.

## Results

Of the 2,567 participants who completed the baseline survey, we excluded 401 participants who did not have any social media accounts and 525 who reported being aged 16 or over. A total of 1,641 participants were included at baseline, of which 551 answered the follow-up survey (33.6%). Descriptive statistics for the weighted sample are reported in Table 1. Supplementary Table S1 reports the non-weighted descriptive statistics. Participants were on average 14.0 years old and 56.5% were girls. A total of 14.8% (95% CI: 11.9%; 20.0%) of participants were considered as ban adherers according to the strict definition, while 41.5% (95% CI: 36.9%; 46.0%) were considered as ban adherers according to the less stringent definition. Group balance was achieved for most covariates (SMD ≤ .19), except non-binary adolescents (SMD=.37) and language (SMD=.26). Table 2 reports descriptive statistics for study outcomes.

**Table 1.** Weighted descriptive statistics of the sample’s covariates.

| **Variables** | **Whole sample** | **No adherence** | **Adherence** | **SMD** |
|---|---|---|---|---|
|  | 100% | 84.2% | 15.8% | - |
| Baseline covariates |  |  |  |  |
| Age<sup>1</sup> | 14.0 | 14.0 | 14.0 | .08 |
| Gender<sup>2</sup> |  |  |  |  |
| Girls | 56.5% | 56.0% | 59.2% | Ref. |
| Boys | 41.0% | 42.1% | 35.5% | .11 |
| Other | 2.5% | 1.9% | 5.3% | .37 |
| Language<sup>2</sup> |  |  |  |  |
| Other | 13.4% | 14.7% | 6.1% | Ref. |
| English | 86.6% | 85.3% | 93.9% | .26 |
| SEIFA score<sup>1</sup> | 975.8 | 976.6 | 971.7 | .09 |
| Time spent on social networks<sup>1</sup> | 20.4 | 19.9 | 23.4 | .19 |
| Bullying<sup>2</sup> |  |  |  |  |
| No | 39.4% | 40.0% | 36.0% | Ref. |
| Yes | 60.6% | 60.0% | 64.0% | .08 |
| RCADS-11 total score<sup>1</sup> | 9.0 | 8.8 | 9.9 | .15 |
| RCADS-11 impact subscore<sup>1</sup> | 10.6 | 10.4 | 11.7 | .16 |
| CHU9D utility score<sup>1</sup> | 0.7 | 0.7 | 0.7 | .18 |
| Score of CHU9D activity-related items<sup>1</sup> | 5.2 | 5.2 | 5.5 | .15 |
| Academic self-efficacy<sup>1</sup> | 23.4 | 23.3 | 24.1 | .12 |
| Exposure |  |  |  |  |
| Adherence to the ban (strict definition)<sup>2</sup> |  |  |  |  |
| No | 84.2% | 100% | 0% | - |
| Yes | 15.8% | 0% | 100% | - |
| Adherence to the ban (less stringent definition)<sup>2</sup> |  |  |  |  |
| No | 58.5% | 69.7% | 0% | - |
| Yes | 41.5% | 30.3% | 100% | - |

SMD: standardised mean/proportion differences; SEIFA: Socio-Economic Indexes for Areas; RCADS: Revised Children’s Anxiety and Depression Scale; CHU9D: Child Health Utility 9 Dimensions.
Weights were constructed using inverse probability of treatment (accounting for differences between adherers and non-adherers) and attrition weights (accounting for baseline differences between completers and non-completers), with imputation (imputation by chained equation with m=1) for baseline missing covariates. The total number of participants may differ slightly from the total sample size due to weight truncation.
<sup>1</sup> Means and absolute standardised mean differences for covariates are reported.
<sup>2</sup> Percentages and absolute standardized proportion differences for covariates are reported.

**Table 2.** Descriptive statistics of the sample’s outcomes.

| **Variables** | **Whole sample** | **No adherence** | **Adherence** | **Missing values** |
|---|---|---|---|---|
| RCADS-11 total score | 8.6 | 8.9 | 8.6 | 14 |
| RCADS-11 impact subscore | 10.2 | 10.5 | 10.2 | 14 |
| CHU9D utility score | 0.7 | 0.7 | 0.7 | 32 |
| Score of CHU9D activity-related items | 5.1 | 5.2 | 5.1 | 32 |
| Academic self-efficacy | 22.6 | 22.3 | 22.6 | 46 |

RCADS: Revised Children’s Anxiety and Depression Scale; CHU9D: Child Health Utility 9 dimensions.
Means are reported.

The results of the analyses of the effect of strict adherence to the ban on the study outcomes are presented in Table 3. The ban had no statistically significant effect on the RCADS-11 total score (p=.321) or the RCADS-11 impact subscore (p=.398). There was a significant effect on the CHU9D utility score (b=0.11, p=.002), the score of CHU9D activity-related items (b=-1.09, p < .001), and academic self-efficacy (b=2.27, p=.033). Overall, those who adhered to the ban experienced better functioning (higher utility, lower impact on daily functioning, and higher academic self-efficacy). The results of the effect of a less stringent definition of ban adherence on the study outcomes are reported in Supplementary Table S2. No effect was statistically significant (p ≥ .244).

**Table 3.** Effect of the strict ban adherence on study outcomes.

| **Outcome** | **n** | **Coefficient** | **Robust s.e.** | **p** | **95% CI** | **R<sup>2</sup>** |
|---|---|---|---|---|---|---|
| RCADS-11 total score | 537 | -1.13 | 1.14 | .321 | -3.37; 1.10 | 30.7% |
| RCADS-11 impact subscore | 537 | -1.20 | 1.41 | .398 | -3.97; 1.58 | 31.0% |
| CHU9D utility score | 519 | 0.11 | 0.04 | .002 | 0.04; 0.18 | 30.5% |
| Score of CHU9D activity-related items | 519 | -1.09 | 0.28 | <.001 | -1.64; -0.54 | 22.7% |
| Academic self-efficacy | 505 | 2.27 | 1.06 | .033 | 0.18; 4.37 | 23.6% |

RCADS-11: Revised Children’s Anxiety and Depression Scale; CHU9D: Child Health Utility 9 dimensions; s.e.: standard error; CI: confidence interval.
Coefficients are adjusted slopes (reference group: non-adherence to the ban). All models used stabilised truncated inverse probability of treatment and attrition weights with robust standard errors and imputation for baseline covariates, adjusting for age, gender, language, Socio-Economic Indexes for Areas score, log-transformed hours of social media use, bullying, pre-intervention RCADS-11 total score, and the baseline level of the outcome.

## Discussion

This study investigated whether adherence to the Australian social media ban for adolescents under the age of 16 impacted their psychological distress, HRQoL, and daily functioning. We employed an emulated trial to address confounding and selection biases, enabling us to draw causal inferences regarding the benefits of the social media ban (19).

### Summary of main findings

Under the strict definition of adherence, the results for the primary outcome suggested that the social media ban had no effect on psychological distress, as assessed using the RCADS-11 three months after the ban. This finding was consistent with previous results from studies of smartphone bans, which have shown limited effects on psychological distress and mental health (e.g., with Australian adolescents, 16, 23). Two previous meta-analyses focusing on the impact of social media restrictions on well-being concluded that there were small benefits that our study was not powered to detect (7, 8).

The results for the secondary outcomes showed that the ban could impact HRQoL and daily functioning. We identified statistically significant associations between strict adherence to the ban and the CHU9D utility score, the score of CHU9D activity-related items, and academic self-efficacy. CHU9D utility was significantly higher in ban adherers than in non-adherers, indicating that overall HRQoL was higher in adolescents who adhered to the ban. Similarly, subscores based on questions related to daily life functioning (e.g., schoolwork/homework, daily routine, and activities) and academic achievement were significantly better in adherers than in non-adherers. Therefore, adolescents who strictly adhered to the ban might have experienced fewer disruptions to their daily lives, leading to better functioning across several domains (24). The magnitude of the results was small for the score of CHU9D activity-related items (decrease of 1.09 point on average, on a scale ranging from 3 to 15) and academic self-efficacy (increase of 2.27 points on average, on a scale ranging from 7 to 35). However, small effects can still be relevant from a public health perspective. When applied to a whole population of adolescents, modest improvements at an individual level can result in large gains at a population level (25). The CHU9D utility score showed an average improvement of 0.11, which was higher than the minimally important CHU9D difference of 0.03 (26, 27). These results are consistent with the findings identifying the areas in which problematic social media use has been shown to interfere with the lives of adolescents, including everyday activities, schooling, and sleep (2, 4). They also support the idea that daily functioning should be considered alongside psychological distress and mental health when evaluating such policies and that it should not be relegated to a secondary outcome.

There are two plausible mechanisms that could explain the impact of adherence to the ban on daily functioning. The first is time displacement: time spent on social media is likely to reduce the time allocated to activities that support daily functioning, such as sleep and schoolwork. Therefore, reducing social media use frees up time for other daily activities (28).

Time displacement due to digital activities has already been described and can be assessed using the “red box, green box” approach (29). The ‘red box’ refers to time spent on digital activities that should be spent doing something else (29). Reducing ‘red box’ hours may therefore improve daily functioning. A second mechanism is reduced attentional interference. For example, social media use may interfere with attention during class and homework (30). However, the evidence regarding the impact of social media on attention remains inconclusive, and further studies with robust designs are needed to investigate this potential mechanism (31).

Conversely, no statistically significant association was identified for the impact subscore of the RCADS-11, which was calculated using two items that assessed the impact of depression- and anxiety-related difficulties on daily life. Given we found no effect of the ban on anxiety and depressive symptoms (RCADS-11 total score), the absence of an effect on impairment attributable to symptoms was coherent.

### The importance of adherence

Our study showed that, when using a strict definition of adherence based on self-reported adherence plus additional variables assessing self-reported behaviours on social media (i.e., reporting sharing/posting on social media, reporting use of social media during free time and indicating regular use of social media accounts targeted by the ban), only 14.8% (95% CI: 11.9%; 20.0%) of adolescents adhered to the social media ban. This figure is consistent with that of a previous study which also reported low adherence: 85% of Australian adolescents continued to use social media after the ban (9). Using a less stringent definition based on self-reported adherence only, 41.5% (95% CI: 36.9%; 46.0%) of adolescents were categorised as adhering to the ban. The discrepancy between these two definitions may be due to social desirability, recall bias, or misunderstanding of the ban (32).

Sensitivity analyses using a less stringent definition of adherence showed no significant effect on the study outcomes. These results may be due to measurement error, as a single question may not accurately capture adherence. If adherers continued to use social media, this would result in misclassification and bias estimates towards the null. Another possible explanation is that benefits only emerge with complete disengagement, as the strict definition captured adolescents who had fully stopped using social media.

Overall, our results highlighted that adherence is a critical factor to assess the potential impact of digital interventions such as a national social media ban. Previous studies have shown that adherence to such interventions is generally low (9, 32). Future interventions should therefore be designed with the aim of maximising adherence to achieve the desired effects, such as psychoeducation about the benefits for young people and parents, help kits to help parents managing their children’s social media use, or federal government efforts to engage with big tech companies to enforce stricter regulations. In our study, an adherence rate of 14.8% three months after implementation suggests that the ban will have a limited population-level impact, despite potential individual-level benefits.

### Limitations

This study had several limitations. First, it relied on self-reported measures. Our adherence to the intervention was also assessed using a self-reported measure and was therefore likely influenced by social desirability and recall biases. Adherence is a key factor in assessing the feasibility of interventions and correctly interpreting their effectiveness (32). Future studies should rely on objective measures of social media use to better assess adherence. Furthermore, the methodological approach to analysing such interventions should include not only intention-to-treat analyses, which miss potential dose-response effects, but also analyses that account for adherence levels. Second, the study was only powered to detect medium effect sizes. Previous meta-analyses have reported small effect sizes for the impact of social media restrictions on psychological well-being (7, 8). Our study may have missed such small effects. This is an important limitation, given that small effects are not necessarily trivial (33, 34). Third, the attrition rate of the study was high, with only 33.6% of participants completing the follow-up. Although IPAW helped to reduce selection bias, it cannot take unmeasured confounders into account, thereby limiting the generalisability of our study. In addition, balance was not achieved for two covariates. Data were also collected in 13 of the 57 invited schools in a single Australian state, which further limits generalisability, but speaks to the practical difficulties in recruiting from school populations. Fourth, the post-intervention survey took place three months after the social media ban was introduced. Consequently, our study only captured the short-term effects on the outcomes of the study. To better understand the potential benefits of the social media ban, future studies should investigate how these outcomes evolve over time.

### Conclusion

Overall, our findings suggested that nationwide bans on social media use could lead to small short-term benefits in HRQoL and daily functioning of adolescents, with no evidence of an effect on psychological distress. However, this only applied when comparing individuals who had genuinely disengaged from social media with those who were still using it. Therefore, ensuring adherence appears to be a key challenge for such policies. Given the limited population-level impact evidenced in the present study, we call for industry-level obligations placing responsibility on platforms to implement “safety by design” principles, coupled with further parental and youth psychoeducation.

## Acknowledgments

This research was supported by a grant from the Channel 7 Children’s Research Foundation. DLK is supported by a 2024 National Health and Medical Research Council (NHMRC) Investigator L1 Fellowship. The funders of the study did not participate in the study design, data collection, analysis or interpretation, or the writing of the report, nor did they influence the decision to submit the paper for publication.

## Data sharing statement

The de-identified individual participant data that underlie the results reported in this article and Stata code will be made available upon reasonable request to the corresponding author from the time of publication onwards. The data will be shared for replication purposes only, with the approval of the last author. This is in accordance with the conditions of the original ethics approval. The pre-registration of the study and questionnaire are publicly available on the Open Science Framework (https://osf.io/q2tb9).

## Declaration of generative AI and AI-assisted technologies in the manuscript preparation process

During the preparation of this work, the first author used DeepL Write to assist with language refinement and to improve readability. After having used this tool, all authors reviewed and edited the content as needed and take full responsibility for the content of the published article.

---

**Supplementary Table S1.** Unweighted descriptive statistics of the sample, without imputation.

| **Variables** | **Whole sample** | **No adherence** | **Adherence** | **Missing values** | **SMD** |
|---|---|---|---|---|---|
| n<sup>1</sup> | 551 | 85.8% (471) | 14.2% (78) | - | - |
| Baseline covariates |  |  |  |  |  |
| Age<sup>2</sup> | 14.1 (0.9) | 14.1 (0.9) | 13.7 (0.8) | 8 | .46 |
| Gender<sup>1</sup> |  |  |  | 11 |  |
| Girls | 53.1% (287) | 53.7% (249) | 50.0% (37) |  | Ref. |
| Boys | 44.8% (242) | 44.4% (206) | 47.3% (35) |  | .08 |
| Other | 2.0% (11) | 1.9% (9) | 2.7% (2) |  | .05 |
| Language<sup>1</sup> |  |  |  | 9 |  |
| Other | 15.1% (82) | 16.1% (75) | 9.5% (7) |  | Ref. |
| English | 84.9% (460) | 83.9% (391) | 90.5% (67) |  | .21 |
| SEIFA score<sup>2</sup> | 978.2 (56.9) | 977.0 (57.8) | 984.6 (51.2) | 67 | .15 |
| Time spent on social networks<sup>2</sup> | 19.6 (19.8) | 20.3 (20.2) | 14.5 (15.7) | 40 | .33 |
| Bullying<sup>1</sup> |  |  |  | 93 |  |
| No | 40.6% (186) | 39.8% (157) | 45.3% (29) |  | Ref. |
| Yes | 59.4% (272) | 60.2% (237) | 54.7% (35) |  | .09 |
| RCADS total<sup>2</sup> | 8.7 (7.4) | 9.0 (7.5) | 7.2 (6.5) | 72 | .20 |
| RCADS impact subscale<sup>2</sup> | 10.3 (8.7) | 10.6 (8.8) | 8.6 (7.9) | 72 | .19 |
| CHU9D utility<sup>2</sup> | 0.8 (0.2) | 0.7 (0.3) | 0.7 (0.3) | 83 | .22 |
| Score of CHU9D activity-related items<sup>2</sup> | 5.1 (2.5) | 5.2 (2.5) | 4.7 (2.2) | 82 | .18 |
| Academic self-efficacy questionnaire<sup>2</sup> | 23.1 (6.8) | 22.9 (6.8) | 24.6 (6.6) | 100 | .22 |
| Exposure |  |  |  |  |  |
| Adherence to the ban (strict definition)<sup>1</sup> |  |  |  | 2 |  |
| No | 85.6% (470) | 100% (471) | 0% (0) |  | - |
| Yes | 14.4% (79) | 0% (0) | 100% (78) |  | - |
| Adherence to the ban (less stringent definition)<sup>1</sup> |  |  |  | 9 |  |
| No | 60.0% (325) | 70.0% (325) | 0% (0) |  | - |
| Yes | 40.0% (217) | 30.0% (139) | 100% (78) |  | - |
| Outcomes |  |  |  |  |  |
| RCADS total<sup>2</sup> | 8.6 (7.3) | 9.0 (7.4) | 6.5 (6.5) | 14 | - |
| RCADS impact subscale<sup>2</sup> | 10.2 (8.6) | 10.6 (8.7) | 7.6 (7.9) | 14 | - |
| CHU9D utility<sup>2</sup> | 0.8 (0.2) | 0.7 (0.3) | 0.8 (0.2) | 32 | - |
| Score of CHU9D activity-related items<sup>2</sup> | 5.1 (2.4) | 5.2 (2.5) | 4.2 (1.5) | 32 | - |
| Academic self-efficacy questionnaire<sup>2</sup> | 22.6 (7.3) | 22.3 (7.2) | 25.0 (7.5) | 46 | - |

SMD: standardised mean/proportion differences; SEIFA: Socio-Economic Indexes for Areas; RCADS: Revised Children's Anxiety and Depression Scale; CHU9D: Child Health Utility 9 dimensions.
<sup>1</sup> Percentages, n, and absolute standardised proportion differences for covariates are reported.
<sup>2</sup> Means, standard deviations, and absolute standardised mean differences for covariates are reported.

**Supplementary Table S2.** Effect of the less stringent ban adherence on study outcomes.

| **Outcome** | **n** | **Coefficient** | **Robust s.e.** | **p** | **95% CI** | **R<sup>2</sup>** |
|---|---|---|---|---|---|---|
| RCADS-11 total score | 537 | 0.51 | 0.65 | .434 | -0.77; 1.78 | 32.4% |
| RCADS-11 impact subscore | 537 | 0.65 | 0.79 | .415 | -0.91; 2.20 | 32.6% |
| CHU9D utility score | 519 | 0.01 | 0.02 | .727 | -0.04; 0.05 | 33.6% |
| Score of CHU9D activity-related items | 519 | -0.07 | 0.20 | .741 | -0.47; 0.33 | 26.1% |
| Academic self-efficacy | 505 | 0.76 | 0.65 | .244 | -0.52; 2.03 | 23.8% |

RCADS-11: Revised Children’s Anxiety and Depression Scale; CHU9D: Child Health Utility 9 dimensions; s.e.: standard error; CI: confidence interval.
Coefficients are adjusted slopes (reference group: non-adherence to the ban). All models used stabilised truncated inverse probability of treatment and attrition weights with robust standard errors and imputation for baseline covariates, adjusting for age, gender, language, Socio-Economic Indexes for Areas score, log-transformed hours of social media use, bullying, pre-intervention RCADS-11 total score, and the baseline level of the outcome.

